# Task-related excitatory/inhibitory ratios in the fronto-striatal circuitry predict attention control deficits in attention-deficit/hyperactivity disorder

**DOI:** 10.1101/2021.03.25.21254355

**Authors:** Ping C. Mamiya, Todd L. Richards, Richard A.E. Edden, Adrian K.C. Lee, Mark A. Stein, Patricia K. Kuhl

**Affiliations:** Institute for Learning & Brain Sciences, University of Washington, Seattle, WA, USA; Department of Radiology, University of Washington,Seattle, WA, USA; Department of Radiology and Radiological Science, Johns Hopkins University, Baltimore, MD, USA; Department of Speech and Hearing Sciences, University of Washington, Seattle, WA, USA; Department of Psychiatry and Behavioral Sciences, University of Washington, Seattle, WA USA

**Author notes:** Correspondence to: Ping C. Mamiya, Portage Bay Building, BOX 357988, University of Washington, Seattle, WA 98195 USA.

**Keywords:** Neurotransmitter, Striatum, Basal Ganglia, Executive Function

## Abstract

Reduced GABA concentrations at rest in the fronto-striatal circuitry are repeatedly implicated in cognitive symptoms of ADHD. However, recent evidence has suggested that GABA and its precursor, glutamate, are capable of undergoing dynamic modifications in response to environments. Yet, it remains unclear how the dynamics between glutamate and GABA may change when people are exerting their control of attention, and whether they would predict attention control deficits in ADHD. To study this question, we used MR spectroscopy to quantify GABA and glutamate+glutamine (Glx) concentrations in the anterior cingulate cortex (ACC) and the caudate nucleus in the fronto-striatal circuitry while subjects were performing attention control tasks. We studied 19 adults with ADHD (31-51 years) and 16 adults without ADHD (28-54). We found GABA and Glx concentrations during the tasks increased in both subjects with or without ADHD, but the extent of increases was significantly reduced in subjects with ADHD. Notably, E/I ratios (Glx/GABA) also increased and significantly predicted error rates while subjects with or without ADHD performed the Stroop and Flanker tasks. Critically, regression models including E/I ratios, GABA concentrations, and the ADHD diagnosis significantly predicted task performance in these tasks. Furthermore, clear interactions among these factors predicted the impaired attention control in the Flanker task in subjects with ADHD. These findings demonstrate for the first time that E/I ratios in the ACC and the caudate nucleus increased when people exerted their control of attention, and suggest that reduced GABA contribution to E/I ratio in these two brain regions may account for cognitive deficits in ADHD.

## Introduction

ADHD is a neurodevelopmental disorder that affects approximately 6% of children in the United States (http://cdc.gov/ncbddd/adhd/data.html), and about 5% of children worldwide (Polanczyk *et al*., 2014). Its core symptoms, including inabilities to sustain attention, hyperactivity and impulsivity, often persist into adulthood among affected individuals, leading to lower educational attainment, socio-economic and health crisis (Barbaresi *et al*., 2013; Feldman and Reiff, 2014; Visser *et al*., 2014; Demontis *et al*., 2019). Symptoms in ADHD are thought to result from aberrant development of the fronto-striatal circuitry (Castellanos and Proal, 2012; Friedman and Rapoport, 2015; Mueller *et al*., 2017; Boedhoe *et al*., 2020; Chiang *et al*., 2020). In particular, impair behavioral inhibition and attention control in ADHD may be attributed to reduced GABA concentrations in the fronto-striatal circuitry. Previous studies using high precision MEGA-PRESS (MEshcher-GArwood Point RESolved Spectroscopy), coupled with tissue correction approach has reported decreased GABA concentrations in the anterior cingulate cortex (ACC) and the striatum in the fronto-striatal circuitry in children and adults with ADHD (Ende *et al*., 2016; Puts *et al*., 2020). Yet, reported GABA concentrations are quantified when subjects are at rest. Therefore, it remains unknown whether GABA concentrations would differ between subjects with and without ADHD when they are exerting their control of attention in the tasks, and whether these differences would predict their task performance.

In the mammalian brain, the regulation of GABA concentrations is tightly maintained by the glutamate/GABA-glutamine cycle (Bak *et al*., 2006; Mahmoud *et al*., 2019). Glutamate decarboxylase (GAD) enzyme converts glutamate to GABA, which is to be stored in presynaptic vesicles to be released in neurons (Rae, 2014). This metabolic process is known to be highly dependent on neural activity (Patel *et al*., 2005), which allows the nervous system to exhibit malleability to adapt to environmental changes (Gold and Roth, 1979; Yizhar *et al*., 2011; Tran *et al*., 2019). Thus, if the dynamic balance of glutamate and GABA in the fronto-striatal circuitry is crucial for attention control during tasks, glutamate and GABA concentrations will likely exhibit liable changes in response to task demands. Relatedly, impaired attention control in subjects with ADHD may be attributed to deficits in this dynamic balance between glutamate and GABA.

To study this question, we used MEGA-PRESS to quantify dynamic changes in glutamate/glutamine (Glx) and GABA concentrations in the ACC and the caudate nucleus in the fronto-striatal circuitry while subjects were performing various attention control tasks. The ACC and the caudate nucleus are central to the attention control circuitry in the brain. Glutamate-glutamine (GABA) cycle is important for maintaining proper brain functioning and altered Glx and GABA concentrations in these two brain regions are repeatedly implicated in impaired cognition in neuropsychiatric disorders (Brugger and Howes, 2017; Tebartz van Elst *et al*., 2014; Ajram *et al*., 2017; Nelson and Valakh, 2015; Sohal and Rubenstein, 2019). Here, our aim was two-fold: to investigate whether Glx and GABA concentrations increase during the tasks, and to investigate whether the dynamic balance between Glx and GABA concentrations would enable us to predict different task performance in subjects with or without ADHD. Previous studies have shown that Glx concentrations in the ACC increase when healthy adults and patient with mood disorders perform the attention control task (Taylor *et al*., 2015a; Taylor *et al*., 2015b). Here, we hypothesized that Glx concentrations would increase during the tasks in both subjects with or without ADHD. Based on the existing evidence that GABA concentrations in the ACC increase when people select their choices in a reinforcement learning paradigm (Bezalel *et al*., 2019), we also hypothesized that GABA concentrations in the ACC and the caudate nucleus may increase when our subjects selected their attention between conflicting stimuli. In addition to task-related Glx and GABA increases, we anticipated that these increases would be less in subjects with ADHD. It is because previous studies have shown that genetic variants related to the biosynthesis processes in glutamate and GABA are linked to symptoms in ADHD (Marenco *et al*., 2010; Bruxel *et al*., 2016). Since the biosynthetic steps of glutamate-GABA conversion are dynamically regulated by neural activity (Mahmoud *et al*., 2019; Siucinska, 2019), it is reasonable to argue that Glx and GABA concentrations during the tasks would differ between subjects with and without ADHD, and the dynamic balance of Glx and GABA would be expected to explain their task performance.

## Methods and Materials

### Subjects

The study consisted of 18 subjects with ADHD in the ADHD group and 16 subjects without ADHD in the CONTROL group (Table 1). Subjects with ADHD were recruited by sending the study flyer to patients in the “ Mother First, Father Second” cohorts at the “ Program to Enhance Attention, Regulation, and Learning” clinics at the Seattle Children’s Hospital(Schoenfelder *et al*., 2019). Interested subjects contacted researchers and were screened afterwards. Subjects without ADHD were recruited from the local community and were matched by gender, age, language, educational backgrounds, and employment status with subjects in the ADHD group. All subjects met the following criteria: no history of other mental disorders, neurological impairments, developmental disorders, or hearing loss and right-handed. The average age did not differ between subjects with ADHD (31-51 years) and without ADHD (28-54 years) (*F*_(1,33)_=0.159, *p*=0.692). All experimental procedures were approved by the University of Washington Institutional Review Board and conformed to the ethical principles for research on human subjects from the Declaration of Helsinki, as revised in 2008. All subjects gave written consent to participate in the study.

**Table 1.**
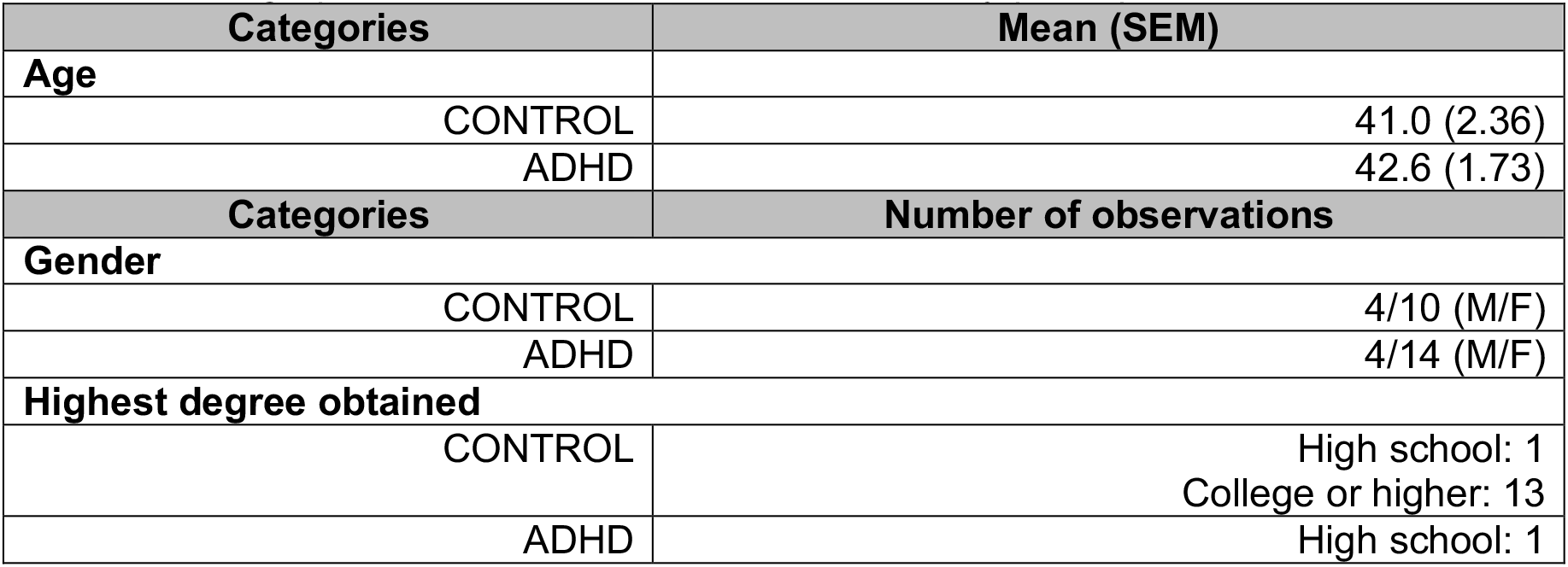
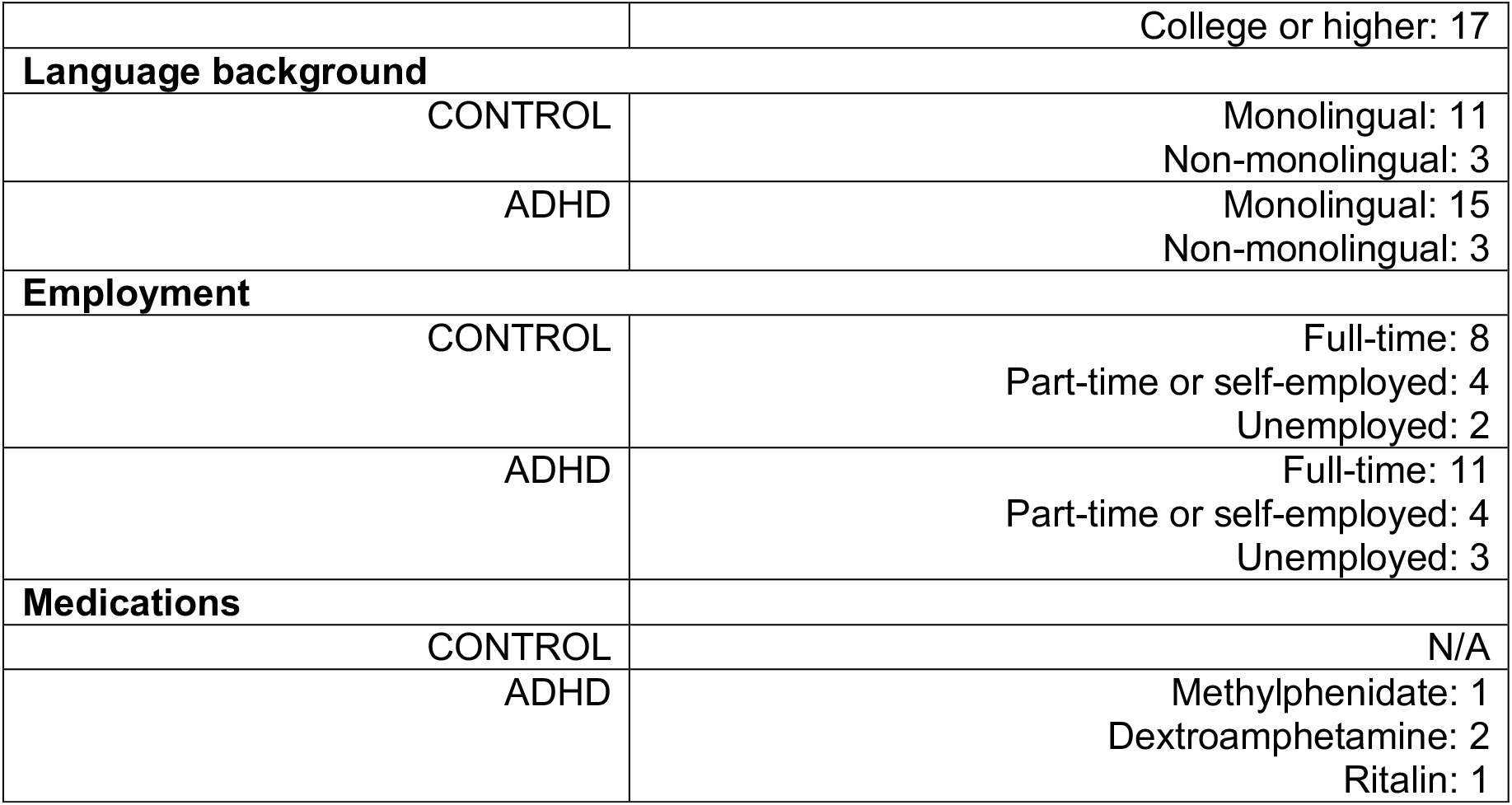
Demographic and clinical characteristics of study participants.

| **Categories** |  | **Mean (SEM)** |
| --- | --- | --- |
| **Age** |  |  |
|  | CONTROL | 41.0 (2.36) |
|  | ADHD | 42.6 (1.73) |
| **Categories** |  | **Number of observations** |
| **Gender** |  |  |
|  | CONTROL | 4/10 (M/F) |
|  | ADHD | 4/14 (M/F) |
| **Highest degree obtained** |  |  |
|  | CONTROL | High school: 1<br>College or higher: 13 |
|  | ADHD | High school: 1 |

|  |  |
| --- | --- |
|  | College or higher: 17 |
| **Language background** |  |
| CONTROL | Monolingual: 11<br>Non-monolingual: 3 |
| ADHD | Monolingual: 15<br>Non-monolingual: 3 |
| **Employment** |  |
| CONTROL | Full-time: 8<br>Part-time or self-employed: 4<br>Unemployed: 2 |
| ADHD | Full-time: 11<br>Part-time or self-employed: 4<br>Unemployed: 3 |
| **Medications** |  |
| CONTROL | N/A |
| ADHD | Methylphenidate: 1<br>Dextroamphetamine: 2<br>Ritalin: 1 |

### Experimental design

All subjects underwent a magnetization-prepared rapid gradient-echo (MPRAGE) T1-weighted imaging and MR spectroscopy (MRS) scans and remained still during the T1 scan. MRS scans consisted of four eight-minutes blocks (Figure 1A). In the first block, all subjects were not required to perform any tasks. In the second block, subjects responded to either low- or high-pitched tones presented in the auditory task. In the third block, subjects viewed various colors of fonts and performed the Stroop task. In the fourth block, subjects viewed a set of conflicting arrows and performed the Flanker task. A MR compatible button-box was placed in each hand (Supplementary FigureS1).

**Figure 1.**
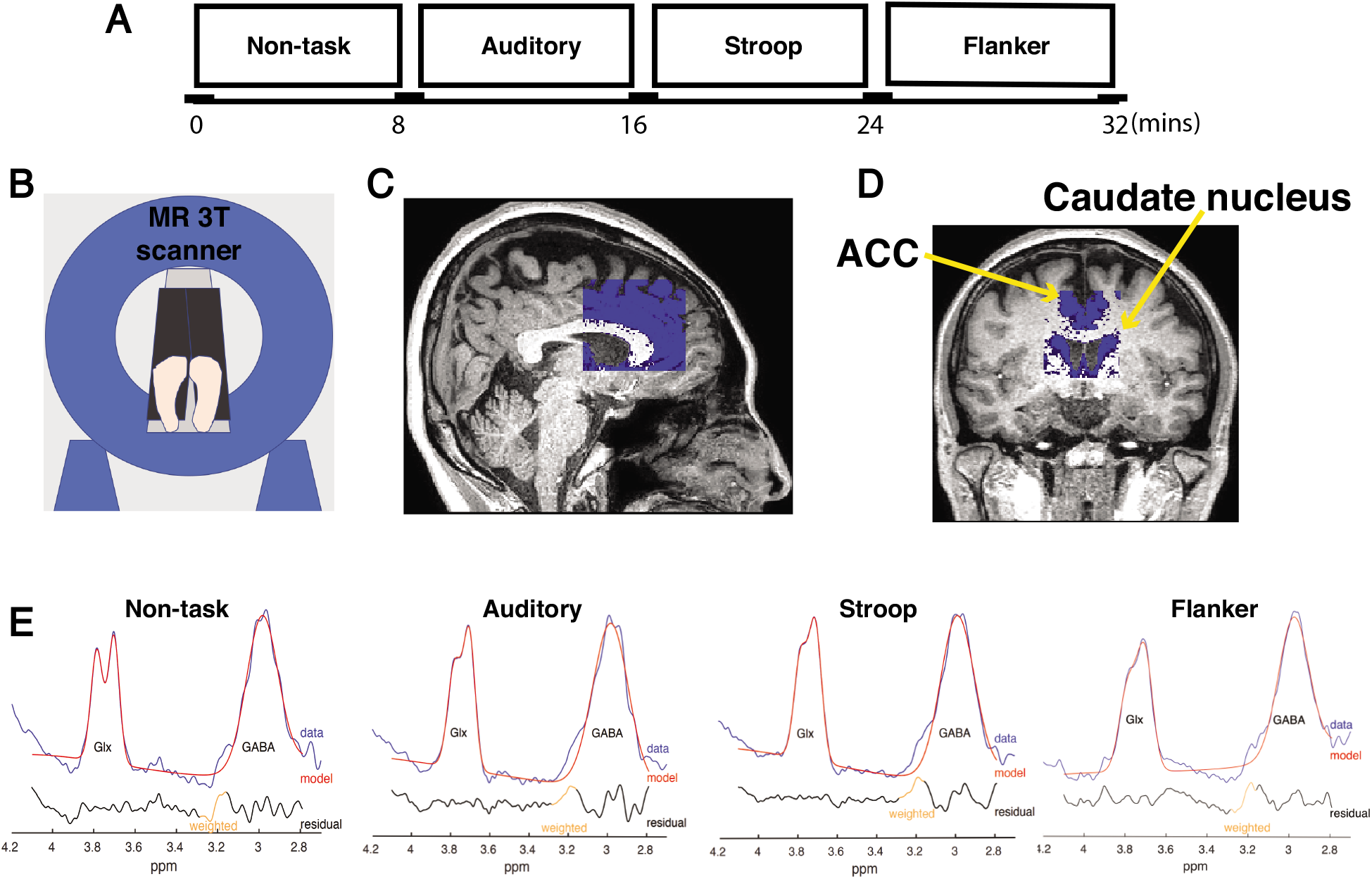
The experimental design, the voxel placement, and the quantification of Glx and GABA concentrations in the MRS experiment. (A) The entire MRS scan was divided into four blocks of time. Each block lasted eight minutes. In the 1^st^ block, all subjects were not required to perform any tasks and remained still insides the scanner. In the subsequent blocks, subjects performed auditory, Stroop and Flanker tasks. (B) A cartoon illustration of a subject laying still inside the Philips Achieva 3T scanner. (C) An example of the voxel placement overlaid on a subject’s T1-weighted image shown in the axial plane. (D) The anterior cingulate cortex (ACC) and the caudate nucleus within the MRS voxels shown in a T1-weighted image in the coronal plane. (E) Representative fits of a single GABA peak at 3 parts per million (ppm) and a double Glx peak at 3.75ppm in the edited MEGA-PRESS spectrum during the non-task and task conditions. The red trace is the model fit overlaid on the raw MRS data.

### Auditory task

We adopted an auditory task that we previously published to assess subjects’ selective attention to auditory stimuli(McLaughlin *et al*., 2019). We used E-Prime to present a 500-ms visual cue (up or down arrow) followed by two spoken digits (high or low pitch) that were played simultaneously during a trial. The pitch of the spoken digits was shifted up and down using Praat software(Boersma and Weenik, 2009) to create competing tokens at 185 Hz ± 4.25 semitones. An arrow cue indicated which spoken digit the subject should be attending to. An up arrow indicated that the target is a high-pitch digit, and a down arrow indicated that the target is a low-pitch digit. Spoken digits were either one, two, three, or four. Subjects were instructed to press the left button in the left hand if they heard the word “ one,” the right button in the left hand for the word “ two,” the left button in the right hand for the word “ three,” and the right button in the right hand for the word “ four”. All subjects completed 70 trials in the task.

### Color-naming version of the Stroop task

We adopted the procedures for the Stroop task that we previously published (Mamiya *et al*., 2018), and used E-Prime to present one of four fonts, GREEN, RED, YELLOW, and BLUE in a mirror inside the scanner. Subjects viewed the color of a font that was consistent with the meaning conveyed by the font in the congruent condition or inconsistent with the meaning of the font in the incongruent condition. We used a single block design and intermixed the congruent and incongruent trials within the block. Subjects were instructed to identify the color of a font in both conditions and use the button-box to respond. The corresponding button for the color of red was the red button, for the color of green the green button, for the color of yellow the yellow button, and for the color of blue the blue button. All subjects completed 186 trials in the task.

### Flanker task

We adopted the procedures for the Flanker task that we previously published (Mamiya *et al*., 2019) and used E-Prime to display 5 arrows in a mirror inside the scanner. Subjects were required to identify whether the arrow of the center image pointed in the same direction (congruent condition) or in the opposite direction (incongruent condition) of the flanking arrows. We used a single block design and intermixed the congruent and incongruent trials within the block. The left button in subjects’ right hand would be pressed if the center arrow pointed to the left. The right button in subjects’ right hand would be pressed if the center arrow pointed to the right. All subjects completed 151 trials in the task.

Subjects were instructed to respond as soon as a stimulus was presented. An invalid trial was marked if there was no button pressed two seconds after a presentation. E-Prime recorded the time when a presentation was shown and the time when a button was pressed in every trial. The latency represented a subjects’ reaction time (RT) in a given trial.

### Magnetic resonance (MR) data acquisition

MR measures were acquired on a Philips 3T Achieva scanner version 5.18 using a 32-channel head coil (Figure 1B). MPRAGE T1-weighted image was acquired using the following parameters: TR=11ms, TE=2.3ms, flip angle=8°, 256 slices covering the entire train, field of view=230×230 mm^2^, matrix size=328×320 mm, reconstructed voxel size= 0.68×0.68×0.70 mm^3^ and was used to place a voxel in each individual’s brain to perform partial volume tissue correction in the subsequent data analysis. The total scan time was four minutes and one second.

GABA-edited MR spectra were acquired using the MEGA-PRESS method (TE=68ms, TR=1,500ms, 1,024 sampling points) with an editing pulse applied either at 1.9 ppm (ON) or at ppm (OFF)(Mescher *et al*., 1998; Edden and Barker, 2007). Water suppression was achieved with a variable power radio frequency pulses with optimized relaxation delays (VAPOR) at the beginning of the MRS scan (Tkac *et al*., 1999). A brain voxel size of 50(anterior-posterior)×40(right-left)×45(foot-head)mm^3^ encompassed the anterior cingulate cortex and the head of the caudate nucleus (Figure 1C,D). The assessment of GABA with MEGA-PRESS included the co-editing of macromolecules, which contributed to the edited peak at 3 parts per million (ppm). Data quality was closely monitored and the acquisition was terminated and restarted if any movement occurred.

### MRS data analyses

We used the Gannet3 toolbox to quantify Glx and GABA concentrations within the voxel(Edden *et al*., 2014). Processing included automatic frequency and phase correction, artifact rejection (frequency correction parameters >3 SD above mean), 3 Hz exponential line broadening, and fitting of the creatine signals. After subtracting OFF from ON acquisitions, a single GABA peak at 3 ppm and a double Glx peak at 3.75 ppm were separately fitted using a five-parameter Gaussian mode. A GABA peak was fit with a Gaussian and the integral of the fit served as the concentration measurement. This GABA value was scaled by the integral of the unsuppressed water peak, fit with a mixed Gaussian-Lorentzian.

The surface area in a Glx peak in the MEGA-PRESS difference spectrum was estimated using a double Gaussian fit, and normalized to water. An MRS voxel for each subject was co-registered to its respective structural image using GannetCoRegister (Harris *et al*., 2015). This produced a binary voxel mask, which was segmented into gray matter, white matter, and cerebrospinal fluid (CSF) probabilistic partial volume maps using the unified tissue segmentation algorithm in SPM12 (Ashburner and Friston, 2005) provided by GannetSegment (Harris *et al*., 2015). The GABA and Glx values were then corrected based on segmented T1-weighted images in each individual. To assess the consistency of data quality, full width half maximum (FWHM) of creatine, fit errors of GABA and Glx peaks were additionally assessed and only spectra with FWHM 20Hz or less, or fit error less than 10% were included in the analysis. We found that one subject with ADHD showed excessive motion during block3 and block4. The Glx and GABA model could not be fit in these blocks. Thus, the MRS data from this subject was not entered in the analysis.

We used the MEGA-PRESS OFF spectra and examined creatine (Cr), N-acetylaspartate (NAA), N-acetylaspartylGlu (NAAG) signals using the LC Model (version 6.3-1L) (Provencher, 1993). Only subjects whose Cramer-Rao lower bounds (CRLB) were 20% or lower were entered in the analysis.

### Statistical analyses

We use the tidyverse (version 1.3.0) and rstatix libraries in *R* (version 3.6.2) for data analysis. We calculated the average, standard deviation, and 95% confidence interval of measurement precision of Glx and GABA concentrations (Supplementary Table S1). We divided Glx by GABA to derive the E/I ratio in each individual block and transformed it to a natural log value. We computed the changes in Glx, GABA and E/I ratios in block2, 3 and 4 with respect to block1. To test the hypothesis that the Glx concentrations increased during tasks, we used the ‘group’ (ADHD versus CONTROL) as the between-subject factor, the ‘block’ (block1-block4) as the within-subject factor, and Glx changes (ΔGlx) as a dependent variable in a repeated measures analysis of variance (ANOVA). We added age as a covariate in the repeated measures analysis of covariance (ANCOVA) where group was a between-subject factor, block was a within-subject factor, GABA changes (ΔGABA) or E/I ratio changes (Δln(E/I ratio)) as dependent variables. We used quantile-quantile plot of ANOVA residuals to assess the normality of distribution. Control group was used as the reference level in the ‘group’ factor. Block 1 was used as the reference level in the ‘block’ factor. We used two-way repeated measures ANOVAs to assess the FWHM of Creatine and the fit errors of Glx and GABA peaks and entered the group as the between-subject factor, block as the within-subject factor, and FWHM or fit error as a dependent variable. We used the non-parametric Kruskal Wallis test to assess tissue components within the MRS voxel, and the Glx and GABA concentrations in the non-task condition (block1) in subjects with and without ADHD. In all analyses, Tukey Honest Significant Differences (HSD) was used as a post hoc test after ANOVA and adjusted *p* value according to the number of comparisons.

We calculated task error rates by dividing the number of incorrect responses by the total number of trials in the congruent and incongruent condition respectively. We found one subject without ADHD and one subject with ADHD showed an error rate three SDs higher than the group average. Thus, their data were removed from the Stroop analysis. We used the non-parametric Kruskal Wallis test to understand behavioral measures in subjects with or without ADHD. We performed two-way ANOVA to understand gender effect on the task performance. We entered gender and group as two between-subject factors. We applied Bonferroni correction and multiplied the resulting *p* values by two to account for two conditions in the task.

To study whether the error rates can be explained by the E/I ratios, GABA concentrations and ADHD diagnosis, we used linear regression and entered GABA, E/I ratio and group (ADHD vs. CONTROL) as predictors in the model. We used ANOVA to understand the model fit and the eta squared to determine the effect size of a predictor. We performed permutation tests with 500 iterations to confirm the chance of obtaining the observed R-squared and *p* values of each model was greater than would be expected by chance (*p*<0.05).

We used Pearson correlation test to assess the relationship between age and GABA concentrations, as well as between GABA and Glx concentrations and applied Bonferroni corrections on resulting *p* values. We used the alpha level of 0.05 in the study.

## Results

### 1. Glx increases during the tasks are reduced in adults with ADHD

To test the hypothesis that glutamate concentrations would increase while subjects performed the attention control tasks, we used MEGA-PRESS to quantify glutamate+glutamine (Glx) concentrations and computed the differences in Glx concentrations between task versus non-task conditions. We found significant Glx increases in both subjects with or without ADHD (two-way repeated measures ANOVA: *F*_(3,126)_=9,820, *p*=7.2×10^−6^, η_p_ ^2^=0.190). Notably, these increases were significantly less in subjects with ADHD than subjects without ADHD, indicated by a significant effect of group on Glx increases (for CONTROL: mean±SEM=12.86±2.09, CI=[17.04, 8.68]; for ADHD: mean±SEM=8.07±1.54, CI=[11.13, 5.01]; *F*_(1,126)_=10.295, *p*=0.002, Figure 2A), but not group-by-block interaction (*F*_(3,126)_=0.707, *p*=0.550). A Tukey HSD post hoc analysis revealed significant Glx increases in block2 while subjects performed the auditory task (adjusted *p*=0.004, 95%CI= [2.934, 20.309]), in block3 when subjects performed the Stroop task (adjusted *p*=2×10^−4^, 95% CI= [5.524, 22.899]), and in block4 when subjects performed the Flanker task (adjusted *p*=5×10^−5^, 95% CI= [-6.774, 24.150]). Relatedly, we found that the Glx concentrations during the non-task condition were not different between subjects with and without ADHD (Kruskal-Wallis test: *H*_(1)_ = 0.015, *p*=0.904).

**Figure 2.**
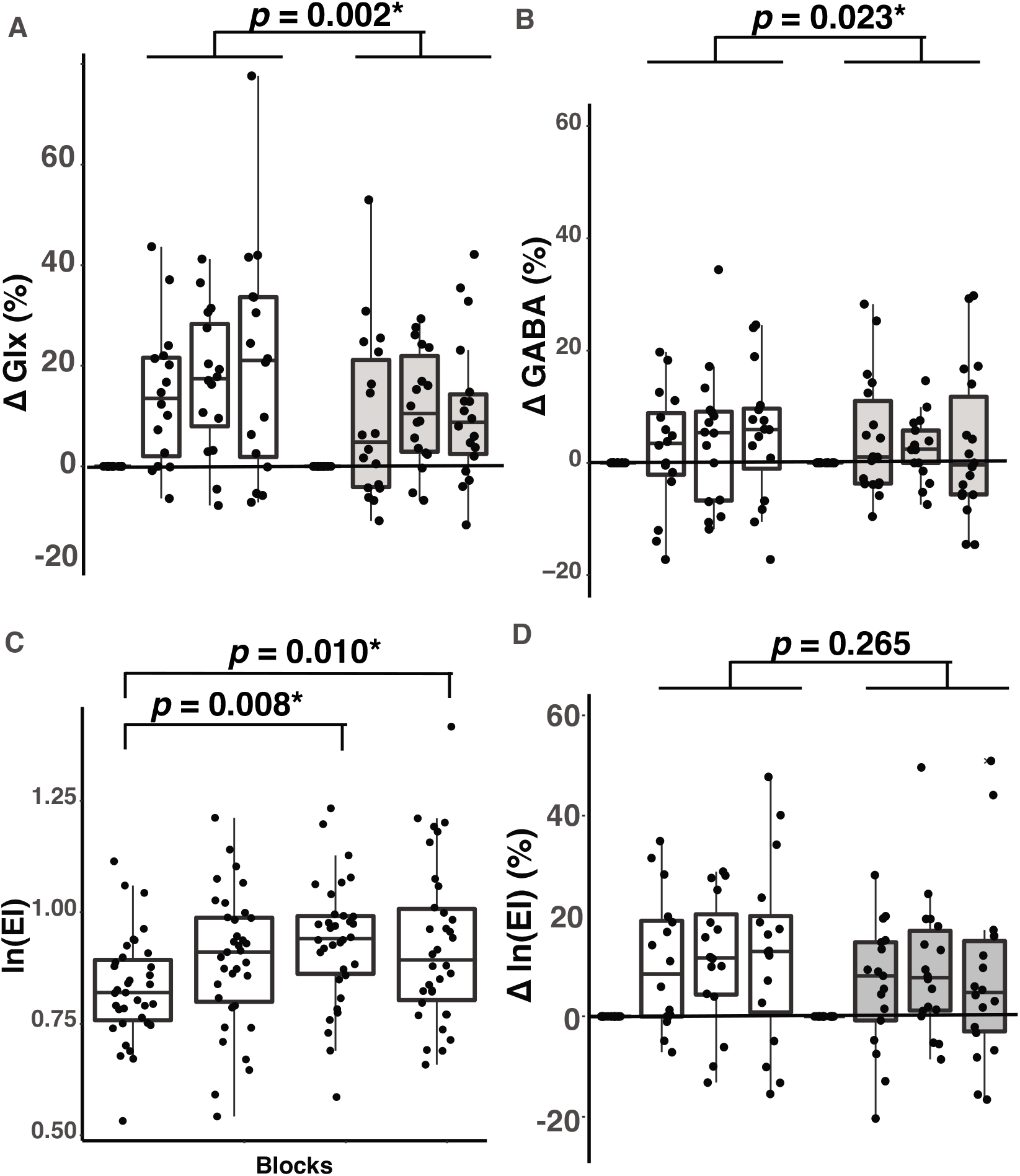
Task-related changes in Glx, GABA, and E/I ratios. Open bars represent subjects without ADHD and grey bars represent subjects with ADHD. (A) Increases in Glx concentrations (ΔGlx) during the tasks are shown in percentage. ΔGlx were computed using the following formula: ΔGlx=(Glx_task_ – Glx_non-task_)/ Glx_non-task_ ×100%. In each group, three bars are shown as the ΔGlx in the auditory, Strroop and Flanker tasks respectively. (B) Increases in GABA (ΔGABA) concentrations are shown in percentage. ΔGABA were computed using the following formula: ΔGABA=(GABA_task_ – GABA_non-task_)/ GABA _non-task_ ×100%. In each group, three bars are shown as the ΔGlx in the auditory, Strroop and Flanker tasks respectively. (C) E/I ratios during the MRS scan. X-axis represent four blocks of time. Subjects did not perform any tasks in block1, but performed the auditory task in block2, Stroop task in block3, and Flanker tasks in block4. Y-axis represents natural log values of E/I ratios (ln(E/I ratio). (D) Increases in E/I ratios (ln(E/I ratio)) during the tasks are shown in percentage. Δln(E/I) were computed using the following formula: Δln(E/I)=(ln(E/I ratio)_task_ – ln(E/I ratio)_non-task_)/ ln(E/I ratio)_non-task_ ×100%. In each group, three bars are shown as the ΔE/I ratios in the auditory, Strroop and Flanker tasks respectively. Dots represent the data points from individual subjects in all figures. The upper boundary of an individual box represents the 75^th^ percentile and the lower boundary represents the 25^th^ percentile of the value for an individual block. The horizontal line within the box represents the median in a respective block.

### 2. GABA concentrations during the tasks are reduced in adults with ADHD

GABA concentrations in the ACC increase when people receive reward based on choices they make in a reinforcement learning paradigm (Bezalel *et al*., 2019). This forced-choice behavior is heavily influenced by brain functions in the ACC and its connected brain regions in the basal ganglia {Dayan, 2008 #2216;Maia, 2011 #2217;Botvinick, 2012 #2218;Holroyd, 2012 #1241}. Our subjects were required to select their attention when facing conflicting stimuli in the attention control tasks. Thus, GABA concentrations in the ACC and the caudate nucleus would increase during the attention control tasks. To study this question, we computed GABA concentrations while subjects were performing various attention control tasks and compared them with GABA concentrations quantified while subjects were at rest. We included age as a covariate due to the strong effect of age on cortical GABA concentrations (Gao *et al*., 2013; Rowland *et al*., 2016). We found that GABA concentrations during the tasks were significantly smaller in subjects with ADHD (mean±SEM=2.91±1.23) compared to subjects without ADHD (mean±SEM=2.23±1.01), indicated by a significant group effect (two-way repeated measures ANCOVA, *F*_(1,125)_=4.651, *p*=0.033, η_p_ ^2^=0.036, Figure 2B), and a trend of increases in both subjects with or without ADHD (block effect: *F*_(3,125)_=1.189, *p*=0.317; group-by-block interaction: *F*_(3,125)_=0.001, *p*=0.968). We additionally assessed whether GABA concentrations at rest differed between subjects with and without ADHD. We did not find GABA concentrations at rest differed significantly (Kruskal-Wallis: H_(1)_ = 0.04, *p*=0.38).

### 3. E/I ratios during the tasks increase in both subjects with or without ADHD

Increases in Glx and GABA concentrations during the tasks are consistent with the existing literature that glutamatergic and GABAergic systems make adaptive changes to environments (Rothman *et al*., 2011); (Hendry and Jones, 1988; Benson *et al*., 1994; Ding *et al*., 1998; Li *et al*., 2010; Cuzon Carlson *et al*., 2011). Yet, animal and human literature has shown mixing results regarding whether glutamate and GABA concentrations would correlate with one another in order to maintain the homeostatic balance of excitation and inhibition when neural activity increases (Xue *et al*., 2014; Bachtiar *et al*., 2015). To study this question, we first examined the relationship between Glx and GABA and then computed the E/I ratios to understand the dynamic balance of Glx and GABA concentrations. We found that Glx concentrations were significantly correlated with GABA concentrations when subjects were at rest in block1 (Pearson *r*=0.564, *p*=0.002), but not correlated GABA concentrations when subjects were performing the tasks (*p*>0.05). Remarkably, we found significant increases in E/I ratios during the task after controlling for age for both subjects with or without ADHD (block effect from a two-way repeated measures ANCOVA: *F*_(3,125)_=4.476, *p*=0.005, η_p_^2^=0.190, Figure 2C), but these increases did not differ between subjects with and without ADHD, indicated by a non-significant group effect (*F*_(1,125)_=1.253, *p*=0.265, Figure 2D), and group-by-block interaction (*F*_(3,125)_=0.435, *p*=0.782). In specific, we found that E/I ratios increased when subjects performed the Stroop task in block3 (Tukey HSD post hoc analysis, adjusted *p*=0.008) and the Flanker task in block4 (adjusted *p*=0.010), but not during the auditory task in block2.

Together, these results suggest that the balance of excitation and inhibition underwent significant increases during the Stroop and Flanker tasks in both subjects with or without ADHD. Notably, both Glx and GABA concentrations during the tasks were significantly reduced in subjects with ADHD compared to subjects without ADHD.

### 4. Glx - GABA relationship at rest are not affected by NAA and NAAG

The observed increases in E/I ratios are in a great agreement with the emerging view that the balance of excitation and inhibition is dynamically regulated in response to environments. However, recent evidence has suggested that N-acetylaspartate (NAA) can affect the balance of excitation and inhibition in the human cortex (Steel *et al*., 2020). Thus, it is possible that observed E/I increases are related to changes in NAA concentrations. To explore this possibility, we studied NAA and its metabolite, N-acetylaspartylGlu (NAAG), and asked whether the relationship between Glx and GABA may be affected by NAA+NAAG, and whether NAA+NAAG increased during the tasks. We found that Glx concentrations were significantly correlated with GABA concentrations after we regressed out NAA+NAAG (linear regression: *R*^*2*^=0.357, *p*=0.001, Supplementary Figure S2A). In addition, NAA+NAAG concentrations did not change during the scan (two-way repeated measures ANOVA: *F*_(3,122)_ = 0.556, *p*=0.645, Supplementary Figure S2B). These findings indicated that the observed E/I increases were attributed to Glx and GABA increases, but not NAA+NAAG concentrations.

### 5. Tissue compositions do not differ between subjects with and without ADHD

The observed reduction in GABA concentrations during the tasks in subjects with ADHD supports the hypothesis that insufficient GABA may contribute to symptoms in ADHD. However, it is known that the proportion of GM in the MRS voxel is crucial for determining Glx and GABA quantification in the MRS analysis, with two times more GABA concentrations in GM than WM (Mikkelsen *et al*., 2016). Thus, we wanted to understand whether the observed smaller Glx and GABA concentrations in subjects with ADHD were attributed to different tissue compositions within the voxel. To answer this question, we studied the fractions of GM, WM and CSF and compared them between the subjects with and without ADHD. We did not find any significant group differences in any of these measurements (Figure 3), indicating that tissue compositions were equivalent between subjects with and without ADHD. Therefore, the fraction of GM composition did not contribute to smaller Glx and GABA concentrations during the tasks in subjects with ADHD.

**Figure 3.**
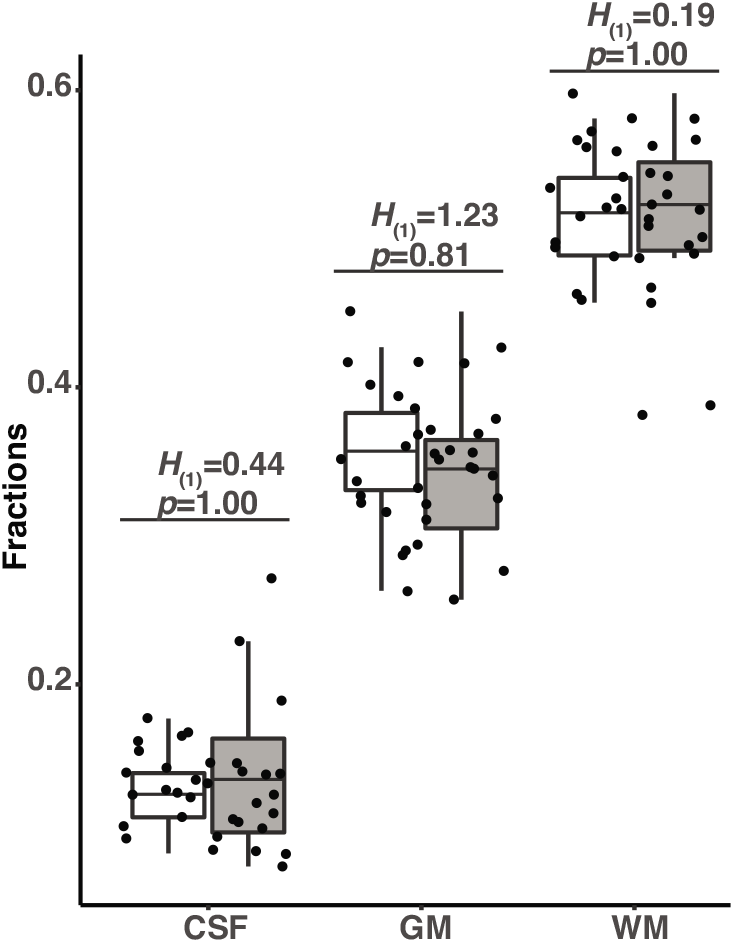
Fractions of gray matter (GM), white matter (WM), and cerebrospinal fluid (CSF) within the MRS brain voxel. Open bars represent subjects without ADHD and grey bars represent subjects with ADHD. Dots represent the data points from individual subjects in all figures. The upper boundary of an individual box represents the 75^th^ percentile and the lower boundary represents the 25^th^ percentile of the value for an individual block. The horizontal line within the box represents the median in a respective block. Statistical results and the corrected *p* values from the Kruskal-Wallis test are shown for GM, WM and CSF respectively.

### 6. Spectral quality is consistent throughout the MRS scan

Another possibility for observing lower Glx and GABA concentrations in subjects with ADHD could be explained by differences in the signal-to-noise ratio (SNR). It has been shown that low SNRs can underestimate Glx and GABA concentrations (Sanaei Nezhad *et al*., 2018) {Steel, 2020 #2219}. To rule out the possibility that reduced Glx and GABA concentrations in subjects with ADHD were due to compromised spectral qualities, we performed two analyses that assessed the linewidth of the creatine signal and the fit errors of Glx and GABA signals respectively. Creatine is a prominent metabolite in the brain and commonly used as a reference signal in the MRS analysis. Thus, the linewidth of the creatine signal and fit errors of Glx and GABA peaks can provide clues of the spectrum quality. We found FWHM of creatine signals to be consistent throughout the scan (two-way repeated measures ANOVA: *F*_(3,128)_=0.042, *p*=0.990, Supplementary Figure S3A). We also found consistent fit errors of Glx peaks throughout the scan (two-way repeated measures ANOVA: *F*_(3,128)_=0.516, *p*=0.672, Supplementary Figure S3B). Similarly, we did not find that the fit errors of GABA peaks changed during the tasks (two-way repeated measures ANOVAs: *F*_(3,128)_=1.207, *p*=0.310) (Supplementary Figure S3C). There results indicate that all subjects showed consistent data quality throughout the scan. Thus, the observed reduction in Glx and GABA concentrations in subjects with ADHD were not likely due to differences in spectral quality.

### 7. Adults with ADHD show significantly higher error rates in the Flanker task

Observed increased in E/I ratios support the current view that the E/I balance is dynamically regulated and alterations in E/I ratios impair behavioral and cognitive functions in neurodevelopmental disorders (Goel and Portera-Cailliau, 2019). To understand the contribution of E/I ratio to attention control deficits in ADHD, we first examined the task performance, and then asked whether E/I ratios predict differences in task performance. Consistent with existing literature, we found that subjects with ADHD showed a significantly higher error rate in the congruent condition (for CONTROL: mean±SEM=0.23±0.17, for ADHD: mean±SEM=1.58±0.58; Kruskal-Wallis test: *H*_(1)_=4.062, *p*=0.04; Figure 4A), and a marginally higher rate in the incongruent condition (for CONTROL: mean±SEM=0.71±0.35, for ADHD: mean±SEM=2.12±0.62; Kruskal-Wallis test: *H*_(1)_=3.114, *p*=0.070) in the Flanker task. In the Stroop task, subjects with ADHD also showed a trend of higher error rate in the incongruent condition (for CONTROL: mean±SEM=11.23±2.02, for ADHD: mean±SEM=15.15±2.44; Kruskal-Wallis test: *H*_(1)_=1.220, *p*=0.269; Figure 4B). Subjects with ADHD did not show significant differences in choosing high- or low-pitched target digits compared to subjects without ADHD in the auditory task (for ADHD: mean±SEM=42.9±2.92, for CONTROL: mean±SEM=53.8±3.33; Kruskal-Wallis test: *H*_(1)_=0.57, *p*=0.448).

**Figure 4.**
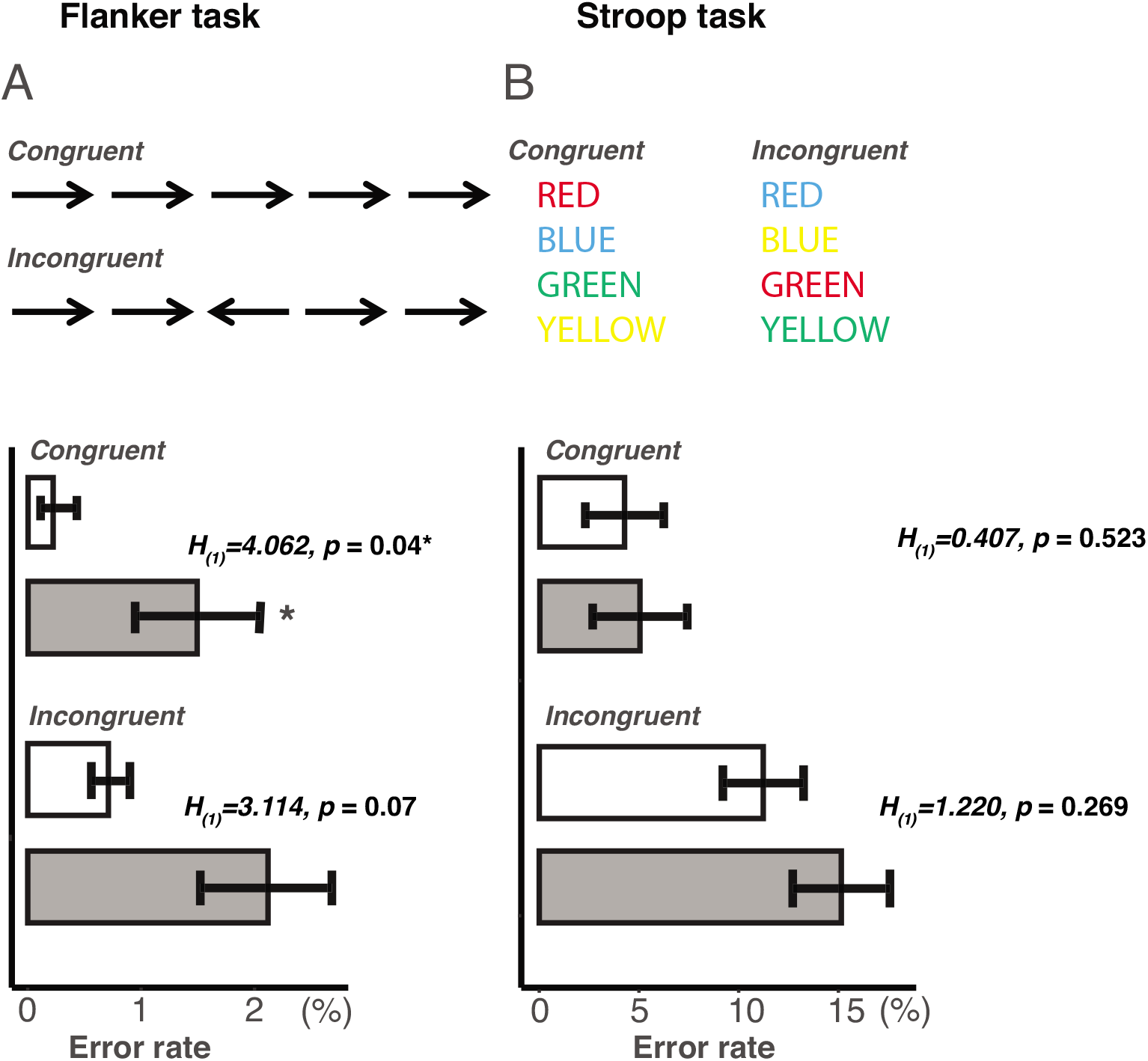
Error rates in the Stroop and Flanker tasks. Each task has two conditions, congruent versus incongruent. Subjects were required to identify the arrow direction in the center image in the Flanker task and identify font colors in the Stroop task. Open bars represent subjects without ADHD and grey bars represent subjects with ADHD. Data is presented as mean with standard error bar. An asterisk indicates a significant difference at a *p* level of 0.05.

### 8. Task-related GABA concentrations, E/I ratios, and ADHD diagnosis predict impaired attention control in subjects with ADHD

Next, we wanted to understand whether the difference in the attention task performances were attributed to E/I ratios, GABA concentrations, and ADHD diagnosis. We used linear regression and found that a linear model consisting of the E/I ratios, GABA concentrations during the Stroop task in block3, and the group variable (ADHD vs. CONTROL) significantly predicted the error rate and explained 24.3% of the total variance in the Stroop task (linear regression: *R*^*2*^=0.243, *F*_(3,28)_=2.994, *p*=0.048, Figure 5A). Importantly, the E/I ratio significantly contributed to the model (ANOVA: *α*=40.43, *F*_(1,28)_=7.309, η_p_^2^=0.194, *p*=0.012), but GABA or the group variable did not (*p*>0.05). To understand how much variance is accounted for by the ADHD diagnosis, we compared the total variance explained by the complete model with the model that consisted of only the E/I ratio and GABA concentrations, and found the total variance decreased from 24.3% to 19.5% (linear regression: *F*_(2,29)_=3.506, *p*=0.043, *R*^*2*^=0.195, Figure 5B), suggesting a sizable variance explained by the ADHD diagnosis. In the Flanker task, we found that a linear model consisting of E/I ratios, GABA concentrations during the Flanker task in block4, and the group variable (ADHD vs. CONTROL) significantly predicted the error rate and explained 52.8% of the total variance (linear regression: *R*^*2*^=0.528, *F*_(7,26)_=4.419, *p*=0.003, Figure 5C). Mainly, GABA concentrations, GABA-by-E/I ratio, and GABA-by-group interactions (ANOVA: *α*=0.325, *F*_(1,25)_=4.978, η_p_^2^=0.09, *p*=0.035 for GABA; *α*=-1.358, *F*_(1,26)_=6.129, η_p_^2^=0.111, *p*=0.020, for GABA-by-EI ratio interaction; *α*=26.498, *F*_(1,26)_ =5.881, η_p_^2^=0.107, *p*=0.023 for GABA-by-group interaction) significantly contributed to the model, suggesting a GABA-driven effect in the Flanker performance. As with the prediction of the Stroop performance, we investigated how much of the total variance can be accounted for by just the GABA concentrations and E/I ratios regardless of the ADHD diagnosis. The model with only GABA concentrations and E/I ratios decreased the total variance explained from 52.8% to 20.1% (linear regression: *F*_(3,30)_=2.51, *p*=0.008, *R*^*2*^=0.201, Figure 5D). We additionally performed permutation tests with 500 iterations and confirmed that the difference between the predicted versus actual error rates was greater than would be expected by chance (*p*<0.05, Supplementary Figure S4). Additional confirmation analysis also revealed that subjects’ gender did not predict error rate in either Stroop or Flanker task (linear regression: Stroop: *F*_(1,27)_=2.530, *p*=0.123; Flanker: *F*_(1,25)_=0.5949, *p*=0.448). For the auditory task, we found that a linear model consisting of the E/I ratio, GABA concentrations during the auditory task and group variables (ADHD vs CONTROL) non-significantly predicted much smaller amount of variability in the task performance (linear regression: *R*^*2*^=0.199, *p*=0.157). Together, these results indicate that GABA concentrations, E/I ratios during the Stroop and Flanker tasks, and the ADHD diagnosis significantly predicted the task performance in subjects with or without ADHD. Importantly, E/I ratio alone is able to predict the performance and interactions among E/I ratios, GABA concentrations and ADHD diagnosis plays a key role in explaining impaired Flanker performance in subjects with ADHD.

**Figure 5.**
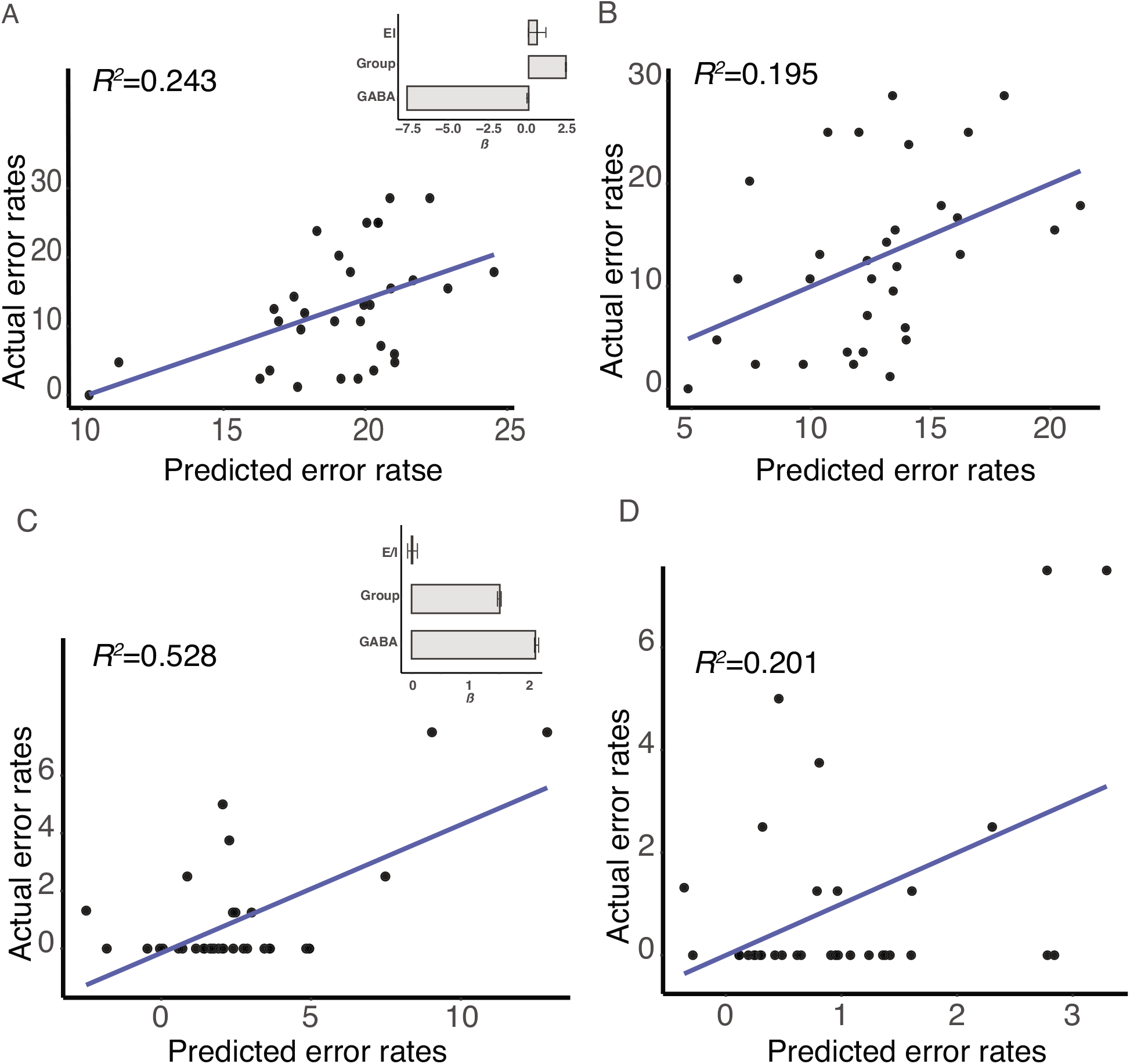
Predictive models for the error rates in the Stroop and Flanker tasks. X-axes represent the predicted error rates derived from the linear models, and y-axes represent the actual error rates. (A) A complete model consisted of the E/I ratio, GABA concentrations in the Stroop task in block3 and the group variable (ADHD vs. CONTROL) significantly predicted the error rate in the Stroop task (*R*^2^=0.243). Insert: the predictors used in the linear regression model and their corresponding regression coefficients (*α*). (B) A model consisted of only the E/I ratio and GABA concentrations in the Stroop task in block3 also significantly predicted the error rate in the Stroop task, but with reduced total variance explained (*R*^2^=0.195). (C) A complete model consisted of the E/I ratio, GABA concentrations in the Flanker task in block4, and the group variable (ADHD vs. CONTROL) significantly predicted the error rate in the Flanker task (*R*^2^=0.528). Insert: the predictors used in the linear regression model and their corresponding regression coefficients (*α*). (D) A model consisted of only the E/I ratio and GABA concentrations in the Flanker task in block4 also significantly predicted the error rate in the Flanker task, but with reduced total variance explained (*R*^2^=0.201).

## Discussion

Difficulties in inhibiting distractions and sustaining attention are hallmark symptoms of ADHD. Children and adults with ADHD have altered Glx and GABA concentrations at rest in the fronto-striatal circuitry, suggesting that the imbalance of excitation and inhibition may explain cognitive impairment in this population. However, Glx and GABA concentrations can change with task demands, and it is unclear how the Glx and GABA concentrations may differ between people with and without ADHD when they perform attention control tasks. Furthermore, it is also unknown whether the dynamic balance of excitation and inhibition enables the prediction of the performance in attention control tasks. Here, we combined MEGA-PRESS and quantitative assessments of attention control to show for the first time that E/I ratios in the ACC and the caudate nucleus in the fronto-striatal circuitry increased when subjects exerted their control of attention, and task-related EI ratios predicted attention control in both subjects with or without ADHD. Remarkably, subjects with ADHD showed reduced task-associated GABA and Glx changes, and a clear interaction between the E/I ratio and GABA in the Flanker task where they made significantly more errors than subjects without ADHD. These findings demonstrate a role of Glx and GABA changes in the fronto-striatal circuitry and a GABA-driven E/I imbalance in ADHD.

### Reduced GABA and Glx concentrations during the tasks suggest altered glutamate-GABA cycling in subjects with ADHD

The present study used MEGA-PRESS GABA-editing pulse to reveal the dynamic balance of Glx and GABA concentrations in the ACC and the caudate nucleus in subjects with or without ADHD during attention control tasks. Existing literature has shown that GABA concentrations at rest are reduced in children and adults with ADHD (Ende *et al*., 2016; Puts *et al*., 2020). Here, we furthered the understanding by showing that GABA concentrations during the attention control were significantly less in subjects with ADHD than subjects without ADHD (Figure 2B). Additionally, we found significant increases in Glx concentrations during the tasks but these increases were also significantly smaller in subjects with ADHD (Figure 2A). These findings suggest that the reduction of GABA and Glx concentrations during the tasks may reflect neurochemical abnormality in people with ADHD.

Reduced Glx and GABA concentrations during the tasks may reflect changes occurred in various steps in the glutamate-GABA cycling, such as synthesis, metabolism and clearance. Increases in Glx and GABA concentrations during the tasks (Figure 2) can reflect increased metabolic activity, and a higher glutamate-GABA cycling rate (Cooper and Jeitner, 2016; Siucinska, 2019) (Hendry and Jones, 1988; Akhtar and Land, 1991; Benson *et al*., 1994; Charpier and Deniau, 1997; Bowers *et al*., 1998; Gierdalski *et al*., 2001; Li *et al*., 2010; Cuzon Carlson *et al*., 2011) that take place in the extracellular space, cytoplasm, and the mitochondria within the voxel (Besse *et al*., 2015) (Rae, 2014; Mahmoud *et al*., 2019; Siucinska, 2019).

Intracellular glutamate – glutamine cycling increases when metabolism increases in the neurons {Hyder, 2013 #2220}. Importantly, glutamate metabolism is implicated in ADHD. Polymorphisms in a gene catalyzing GABA synthesis (glutamate decarboxylase 1, *GAD1*) are associated with hyperactivity and impulsivity symptoms in ADHD (Marenco *et al*., 2010; Bruxel *et al*., 2016). GAD enzyme in neurons catalyzes glutamate – GABA conversion (Sheikh *et al*., 1999). There is evidence that GAD enzymatic activity highly depends on neural activity, and increased GAD activity causes increased GABA conversion in the striatum (Gold and Roth, 1979). In the present study, we did not observe any differences in Glx or GABA concentrations during the non-task condition (block1) between subjects with and without ADHD. Thus, reduced GABA concentrations during the tasks may be related to reduced GAD activity, not a gross reduction in GABAergic neurons in the brain. Further investigations are warrant to characterize whether *GAD1* polymorphisms can affect GABA concentrations during the tasks in people with ADHD.

While altered metabolism in glutamate-GABA cycling may help explain decreased Glx and GABA concentrations in subjects with ADHD, GABA and glutamate clearance in the synaptic clefts could also impact Glx and GABA quantification in the voxel. Once released from neurons, glutamate and GABA transporters uptake these metabolites from the synaptic clefts to astrocytes in order to maintain the homeostasis of excitatory and inhibitory neurotransmission in a local neural circuitry. Both GABA and glutamate transporters are strongly implicated in impaired cognition in neuropsychiatric disorders (Adler *et al*., 2012; Cheng *et al*., 2017; Naaijen *et al*., 2017; Morello *et al*., 2020). It is known that excessive glutamate in the synaptic cleft can cause neurotoxicity, leading to neuronal death (Mahmoud *et al*., 2019). Considering the role of GABA concentrations in sharpening the signal-to-noise ratio, it is not surprising that depletion of GABA transporters can cause hyperactivity and inattention phenotypes in the animal model of ADHD (Chen *et al*., 2015). Consistently, several single-nucleotide polymorphisms (SNPS) in GABA transporter 1 (*GAT1)* gene are associated with symptoms in people with ADHD (Yuan *et al*., 2017). Thus, alterations in GABA and Glx clearance during tasks may be related to reduced GABA and Glx concentrations in subjects with ADHD. Future investigations using animal models of ADHD are needed to verify the association between GABA and Glx transporter bindings and the MRS-based GABA and Glx concentrations.

Our subjects with ADHD showed reduced Glx and GABA concentrations during the tasks in which they showed impaired performance compared to subjects without ADHD. Remarkably, our statistical models demonstrate that the E/I ratio and GABA, but not Glx, concentrations significantly predicted their impaired attention control in the tasks (Figure 5). Our findings aid strongly to the growing body of the literature that task-related GABA concentrations predict task performance(Yoon *et al*., 2016; Ajram *et al*., 2017; Kurcyus *et al*., 2018; Bezalel *et al*., 2019), and highlights the significance of GABA-driven E/I ratios in predicting attention control deficits in ADHD.

Our group analysis revealed that subjects with ADHD had significant higher error rates in the Flanker task compared to subjects without ADHD. The trend of non-significant higher rates in the Stroop task was unexpected given that behavioral studies have repeatedly shown that people with ADHD show impaired performance in this task (King *et al*., 2007; Johnson *et al*., 2008; Herrmann *et al*., 2010; Mullane *et al*., 2011). This may be due to the limitations of the study. First, our attention tasks were presented in a pre-set order. Therefore, the order of the task presented during the scan may have contributed to task performance. In addition, our sample size was modest, allowing us to only provide a medium effect size. Finally, we did not screen for caffeine consumption or smoking in our subjects.

In summary, we combined behavioral assessments of attention control with MRS imaging to demonstrate the dynamics of Glx and GABA concentrations during the attention control tasks in subjects with or without ADHD. E/I ratios, GABA and the ADHD diagnosis predicted attention control in both groups, but GABA interactions with EI ratios predicted the impaired attention control in subjects with ADHD. These findings suggest that the E/I ratio in the fronto-striatal circuitry is related to attention control, and deficiency in GABA signaling in the ACC and the caudate nucleus may contribute to impaired cognition in ADHD.

## Data Availability

Data will be available upon request

## Acknowledgements

This work was supported by The Paros Brain Research Fund (to P.K.K.), NIH R01 EB016089 and P41 EB015909 (to R.A.E.E), NIH R01 DC013260 (to A.K.C.L.), and by NIMH, 1R34MH099208 - 01A1, Seattle Children’s Research Institute, and an investigator-initiated grant from Shire Development LLC, Lexington, MA, a member of the Takeda group of companies (Grant ID IIR-USA-001073) (to M.A.S). We thank Anne B. Arnett for helpful discussions.

## Author contributions

PCM and PKK designed and acquired funding for the experiments. PCM conducted the experiments and performed data analyses; TLR and RAEE developed software for processing and analyzing MRS data; AKCL developed and supervised auditory experiment. MAS provided clinical assessments of ADHD symptoms. All authors contributed to the manuscript preparation.

## Abbreviations

ADHD: Attention-Deficit/Hyperactivity Disorder
ACC: Anterior Cingular Cortex
Glx: glutamate/glutamine
GM: Grey Matter
MEGA-PRESS: MEshcher-GArwood Point RESolved Spectroscopy
MPRAGE: Magnetization -prepared rapid gradient
PFC: Prefrontal Cortex
TE: Echo Time
TR: Repetition Time
WM: White Matter
NAA: acetylaspartate
NAAG: acetylaspartylGlu
FWHM: full width half maximum
CRLB: Cramer-Rao lower bound

