## Supplementary material for "Task-related excitatory/inhibitory ratios in the fronto-striatal circuitry predict attention control deficits in attention-deficit/hyperactivity disorder": ADHD_MRS_SupplementaryMaterials_25March2021.pdf

### Supplementary Information

Figure S1. MR-compatible button box.

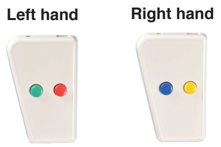


| | CONTROL | | | ADHD | | |
|---|---|---|---|---|---|---|
| | Concentrations | Confidence interval | | Concentrations | Confidence interval | |
| | | Upper | Lower | | Upper | Lower |
| **GABA** | | | | | | |
| Block1 | 2.50 (0.29) | 2.66 | 2.35 | 2.50 (0.32) | 2.66 | 2.35 |
| Block2 | 2.55 (0.29) | 2.71 | 2.40 | 2.63 (0.28) | 2.77 | 2.50 |
| Block3 | 2.59 (0.23) | 2.71 | 2.47 | 2.55 (0.35) | 2.77 | 2.38 |
| Block4 | 2.62 (0.30) | 2.78 | 2.46 | 2.58 (0.32) | 2.74 | 2.43 |
| **Glx** | | | | | | |
| Block1 | 5.84 (0.95) | 6.34 | 5.33 | 5.69 (0.70) | 6.03 | 5.35 |
| Block2 | 6.60 (0.96) | 7.11 | 6.08 | 6.15 (0.73) | 6.51 | 5.80 |
| Block3 | 6.73 (0.63) | 7.06 | 6.39 | 6.35 (0.74) | 6.72 | 5.98 |
| Block4 | 6.90 (1.02) | 7.44 | 6.36 | 6.33 (0.96) | 6.81 | 5.85 |

Supplementary Table S1. GABA and Glx concentrations shown in institution units. Concentrations are shown as mean with standard deviation in the parentheses. Block1: non-task block; Block2: auditory task; Block3: Stroop task; Block4: Flanker task.

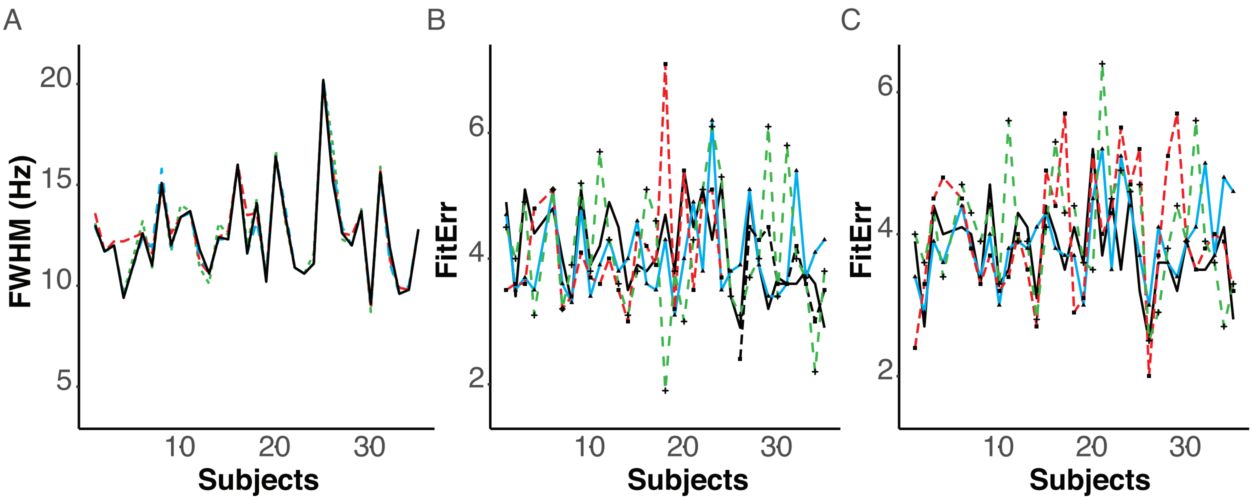


Supplementary Figure S2. MRS quality control measurements. (A) The linewidth of the Creatine signals represented by the full-width of half maximum (FWHM) in four blocks. (B) Fit errors of Glx peaks. (C) Fit errors of GABA peaks. The solid black line represents block1, the green single-dashed line represents block2, the solid blue line represents block3, and the red long dash-line presents block4. X-axis represents study participants and y-axis represents creatine signals shown in Hertz (Hz) for creatine signals or fir errors for Glx and GABA peaks.

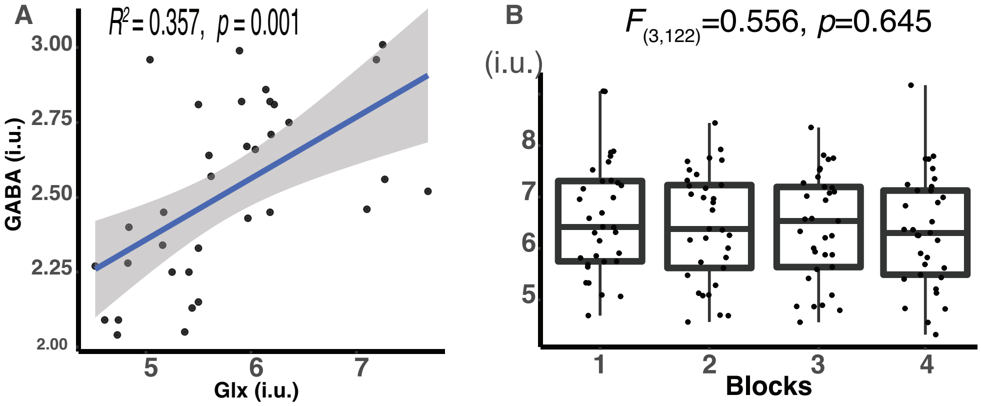


Supplementary Figure S3. (A) A significant relationship between Glx and GABA concentrations when subjects were at rest in block1. Statistical results were obtained by using the linear regression to regress out NAA+NAAG concentrations. X-axis represent Glx concentrations and y-axis represents GABA concentrations. (B) Concentrations of NAA+NAAG/Cr during the scan.

Subjects remained still in block1, and then performed auditory, Stroop and Flanker tasks in block2, block3 and block4 respectively. The upper boundary of an individual box represents the 75th percentile and the lower boundary represents the 25th percentile of the value for an individual block. The horizontal line within the box represents the median in a respective block. In all figures, dots represent the data points from individual subjects.

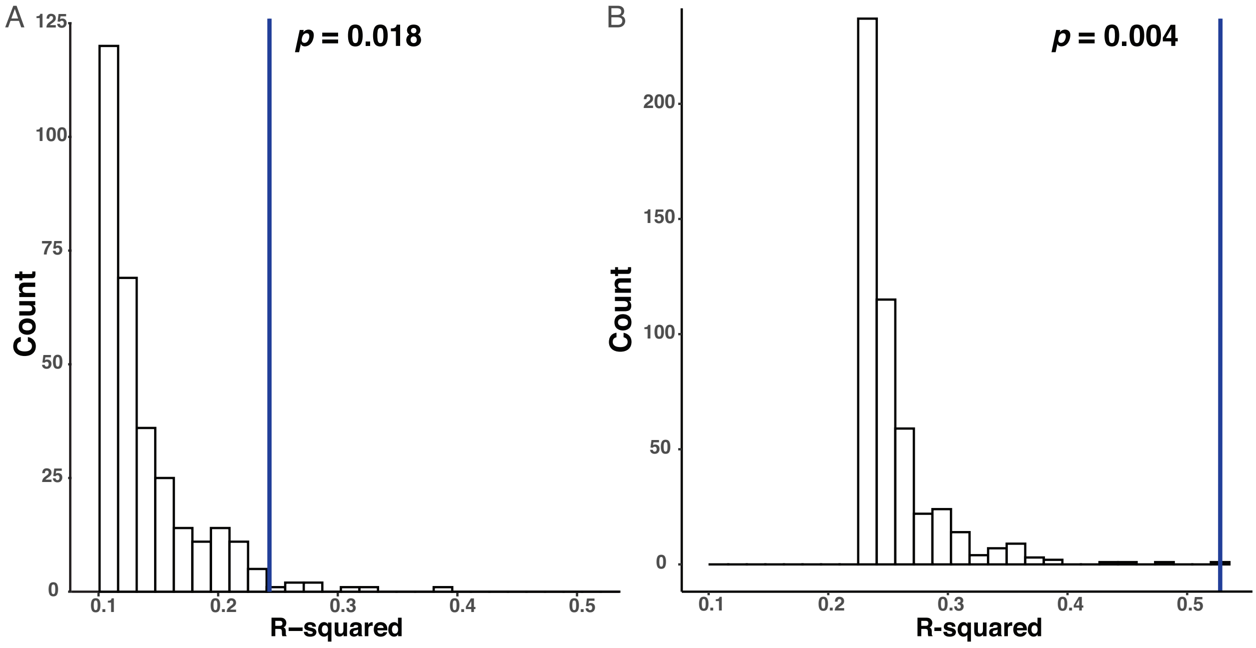


Supplementary Figure 4. Distributions of R-squared populated by 500 permutation tests for (A) Stroop and (B) Flanker tasks. Blue vertical lines represent the R squared in the Stroop task ($R^2$=0.243) and the Flanker task ($R^2$=0.528). *P* values represent the probability of obtaining an R-squared value equal or higher than the observed R-squared value in each model.
